# Genetic susceptibility to acute viral bronchiolitis

**DOI:** 10.1101/2024.02.21.24303021

**Authors:** Anu Pasanen, Minna K. Karjalainen, FinnGen, Matti Korppi, Mikko Hallman, Mika Rämet

## Abstract

Acute viral bronchiolitis is a major cause of infant hospitalizations worldwide. Childhood bronchiolitis is considered a risk factor for asthma, suggesting shared genetic factors and biological pathways. Genetic risk loci may provide new insights into disease pathogenesis. Here, we examined the genetic contributions to bronchiolitis susceptibility by analyzing 1,465 infants hospitalized for bronchiolitis and 356,404 individuals without a history of acute lower respiratory tract infections in the FinnGen project data. Genome-wide analysis identified associations (*p*<5×10^-8^) for variants in gasdermin B (*GSDMB*) and a missense variant in cadherin-related family member 3 (*CDHR3*). Children with bronchiolitis in infancy were more likely to develop asthma later in life compared to controls. The two associated loci were previously linked to asthma and susceptibility to wheezing illness by other causative agents than RSV. Our analysis discovered associations with overall bronchiolitis, with larger effects in non-RSV than RSV-induced infection. Our results suggest that genetic variants in *CDHR3* and *GSDMB* modulate susceptibility to bronchiolitis, especially when caused by viruses other than RSV, and that severe bronchiolitis in infancy may trigger the development of asthma in genetically susceptible individuals determined by these loci.

## Introduction

Acute viral bronchiolitis is a common lower respiratory infection (LRI) that affects infants and young children worldwide. Bronchiolitis is most often caused by respiratory syncytial virus (RSV), followed by other causative viruses such as rhinovirus and metapneumovirus^1–3^. Most infants with bronchiolitis have a mild upper airway disease with cold-like symptoms including a low-grade fever, runny nose, and nasal congestion. Some infants present with severe symptoms (e.g. respiratory distress, fatigue) that require medical attention, and may develop into a condition that is life-threatening requiring intensive care^4^. Bronchiolitis is the leading cause of LRI-related hospitalizations of children younger than two years in developed countries^3,5,6^.

There is no curative treatment for bronchiolitis, and the symptomatic treatment focuses on supporting oxygen supply and maintaining hydration^7^. A maternal RSV vaccine may be given during the third trimester of pregnancy, in the RSV season, to protect the newborn from severe RSV illness^8^. For children at high risk of developing severe RSV bronchiolitis, immunoprophylaxis with the monoclonal antibodies palivizumab or nirsevimab can be used to prevent severe illness^1,9^. Moreover, some infants suffering from rhinovirus bronchiolitis may benefit from systemic corticosteroids^2,7^.

The most important risk factors for severe bronchiolitis include premature birth, chronic lung disease of premature infants, immunodeficiency, and congenital heart disease^3^. However, the known risk factors do not sufficiently explain bronchiolitis severity as most infants hospitalized with bronchiolitis have no identified predisposing conditions. This suggests that genetic and other currently unidentified factors play a role in bronchiolitis severity.

The contribution of genetic variation to bronchiolitis severity is approximately 22%- 40% based on heritability estimates obtained from European twin studies^10,11^. Previous studies of bronchiolitis genetics include several investigations of candidate gene polymorphisms with plausible roles in bronchiolitis pathophysiology^12^. The innate immune response has been shown to affect bronchiolitis severity, and variants in immune system genes have been of particular interest in candidate gene studies of bronchiolitis^13–15^. Variants suggested to be associated with bronchiolitis or RSV bronchiolitis include polymorphisms in genes encoding surfactant proteins (*SFTPA1*, *SFTPA2*, *SFTPD*), *NKG2D*, and *TLR4*^16–19^. Variants suggested to be associated with rhinovirus-induced wheezing or non-RSV bronchiolitis include variants in a genomic region near 17q21 and *CDHR3*^20,21^. Among a few hypothesis-free assessments of bronchiolitis genetics, the GWAS found a suggestive association in *KCND3*^22^. A whole exome sequencing (WES) study suggested associations near genes such as *OR13C5*, *HLA-DQA1,* and *MUC4* with RSV bronchiolitis^23^. Altogether, findings from the genetic studies of bronchiolitis have rarely been replicated in subsequent studies, and the genetic background of bronchiolitis remains largely undercharacterized. A better characterization of host genetic factors can help recognize high-risk infants.

There is a link between severe viral bronchiolitis in infancy and later recurrent wheezing and asthma^1,2,24^. While occasional wheezing symptoms in childhood are common, they may also represent the first manifestation of asthma^25^. Viral bronchiolitis in infancy could activate pathways that lead to the development of asthma^12,26,27^. There are several inflammatory pathways associated with asthma, and multiple asthma-associated genetic loci are known^28,29^. Comparison with bronchiolitis has been limited by the incomplete knowledge of the underlying genetic associations with bronchiolitis. Moreover, recent research suggests more complex interactions between the host, causative respiratory viruses, and subsequent recurrent wheeze or asthmatic symptoms^26,30^. Genetic variants in genes including *TLR4, VDR,* and *CCR5* have been suggested to be associated with both RSV bronchiolitis and the risk of asthma^12,31^. Additionally, genetic variants in genomic region 17q21 and *CDHR3* showed suggestive associations with non-RSV LRI phenotypes and later asthma^20,21,32^.

The aim of the present study was to identify genetic variants that may predispose infants to bronchiolitis by utilizing large-scale genetic data. Moreover, we aimed to assess genetic associations with bronchiolitis linked to RSV and non-RSV and determine the effect of these variants on the risk of developing subsequent asthma.

## Methods

### Ethical considerations

#### FinnGen Ethics statement

Patients and control subjects in FinnGen provided informed consent for biobank research, based on the Finnish Biobank Act. Alternatively, separate research cohorts, collected prior the Finnish Biobank Act came into effect (in September 2013) and start of FinnGen (August 2017), were collected based on study-specific consents and later transferred to the Finnish biobanks after approval by Fimea (Finnish Medicines Agency), the National Supervisory Authority for Welfare and Health. Recruitment protocols followed the biobank protocols were approved by Fimea. The Coordinating Ethics Committee of the Hospital District of Helsinki and Uusimaa (HUS) statement number for the FinnGen study is Nr HUS/990/2017.

The FinnGen study is approved by the Finnish Institute for Health and Welfare (permit numbers: THL/2031/6.02.00/2017, THL/1101/5.05.00/2017, THL/341/6.02.00/2018, THL/2222/6.02.00/2018, THL/283/6.02.00/2019, THL/1721/5.05.00/2019 and THL/1524/5.05.00/2020), Digital and population data service agency (permit numbers: VRK43431/2017-3, VRK/6909/2018-3, VRK/4415/2019-3), the Social Insurance Institution (permit numbers: KELA 58/522/2017, KELA 131/522/2018, KELA 70/522/2019, KELA 98/522/2019, KELA 134/522/2019, KELA 138/522/2019, KELA 2/522/2020, KELA 16/522/2020), Findata permit numbers (THL/2364/14.02/2020, THL/4055/14.06.00/2020, THL/3433/14.06.00/2020, THL/4432/14.06/2020, THL/5189/14.06/2020, THL/5894/14.06.00/2020, THL/6619/14.06.00/2020, THL/209/14.06.00/2021, THL/688/14.06.00/2021, THL/1284/14.06.00/2021, THL/1965/14.06.00/2021, THL/5546/14.02.00/2020, THL/2658/14.06.00/2021, THL/4235/14.06.00/202), Statistics Finland (permit numbers: TK-53-1041-17 and TK/143/07.03.00/2020 (earlier TK-53-90-20) TK/1735/07.03.00/2021, TK/3112/07.03.00/2021) and Finnish Registry for Kidney Diseases permission/extract from the meeting minutes on 4^th^ of July 2019.

The Biobank Access Decisions for FinnGen samples and data utilized in FinnGen Data Freeze 9 include: THL Biobank BB2017_55, BB2017_111, BB2018_19, BB_2018_34, BB_2018_67, BB2018_71, BB2019_7, BB2019_8, BB2019_26, BB2020_1, Finnish Red Cross Blood Service Biobank 7.12.2017, Helsinki Biobank HUS/359/2017, HUS/248/2020, Auria Biobank AB17-5154 and amendment #1 (August 17 2020), AB20-5926 and amendment #1 (April 23 2020) and its modification (Sep 22 2021), Biobank Borealis of Northern Finland_2017_1013, Biobank of Eastern Finland 1186/2018 and amendment 22 § /2020, Finnish Clinical Biobank Tampere MH0004 and amendments (21.02.2020 & 06.10.2020), Central Finland Biobank 1-2017, and Terveystalo Biobank STB 2018001 and amendment 25th Aug 2020.

#### FinnGen dataset

Data from FinnGen Preparatory Phase Data Freeze 9 were used. The Finngen research project (www.finngen.fi) coalesces genome information with digital health care data with help of national personal identification numbers. The GWASs of bronchiolitis and its further sub-groups were conducted with data consisting of 1,456 cases and 356,404 controls.

Phenotypes were defined based on International Classification of Diseases (ICD) entry in the Care Register for Health Care inpatient visits (HILMO) maintained by the Finnish Institute for Health and Welfare (THL). As cases, we included individuals with ICD-10 code J21 for acute bronchiolitis (including J21.0, acute bronchiolitis due to respiratory syncytial virus; J21.8, acute bronchiolitis due to other specified organisms; J21.90, acute obstructive bronchitis of infants; J21.99, unspecified acute bronchiolitis), ICD-9 code 4661A and 46602 for unspecified acute bronchiolitis and bronchitis, and ICD-8 entry 466.99 for acute bronchitis or bronchiolitis. For consistency with bronchiolitis phenotypes in previous studies, the upper age limit was set to two years. All included cases were hospitalized. We further studied bronchiolitis subgroups divided by RSV status using the ICD-10 codes for which RSV infections could be identified. Controls were those without a history of acute lower respiratory infections.

### Genotyping, imputation, and quality control

Genotyping of the FinnGen samples was done with Illumina and Affymetrix arrays (Illumina Inc., San Diego, and Thermo Fisher Scientific, Santa Clara, CA, USA). Eligible samples had non-ambiguous sex, Finnish ancestry, non-high missingness (<5%), and no excess heterozygosity (±4SD). Exclusion criteria in variant quality control (QC) were high missingness (>2%), minor allele count (MAC) <3, and deviation from Hardy-Weinberg equilibrium (HWE, *p*<1e-6). Genotype imputation was done with Beagle 4.1, and a Finnish population-specific SISu reference panel was used. In post-imputation QC, variants with INFO<0.7 were excluded.

### GWAS

We conducted GWAS of bronchiolitis with the FinnGen data with the REGENIE whole-genome regression model implemented in the FinnGen environment. GWAS covariates were age, sex, genotyping batch, and the first 10 principal components. MAC was set to 5 for both cases and controls to be included in the analysis. We conducted a GWAS of bronchiolitis by any causal virus and two subgroup GWA analyses stratified by the RSV status: RSV bronchiolitis GWAS and non-RSV bronchiolitis GWAS.

### Prevalence of asthma after bronchiolitis

We used predefined asthma phenotype and childhood asthma (<16y) to investigate the development of asthma in bronchiolitis cases and controls within the FinnGen data. Asthma definition was based on the diagnosis codes of the Finnish version of the International Statistical Classification of Diseases and Related Health Problems (ICD10:J45, J46; ICD9: 493; ICD8: 493). We used Fisher’s exact test to assess the differences in asthma prevalence between RSV and non-RSV bronchiolitis, and between bronchiolitis and controls.

### eQTL analysis

We used colocalization analysis to assess if cis-eQTL and GWAS signals may share a genetic cause. We used eQTL data from eQTL Catalogue release 6^33^, and Hyprcoloc^34^ to test for colocalization in 500kb windows around the index variants of the associated loci. Suggested by prior knowledge of the bronchiolitis-infected cell types, the analysis was performed in blood, spleen, lung, monocytes, neutrophils, macrophages, B cells, NK cells, LCLs, CD4 T cells, CD8 T cells, T regulatory cells (Tregs), memory Tregs, follicular T helper (Th) cells, Th17 cells, Th1 cells, and Th2 cells.

### pheWas

Previous associations of the lead variants of the associated loci were screened in data from FinnGen DF10.

### Linkage disequilibrium score regression analysis

We used linkage disequilibrium score regression (LDSC) to estimate heritability and genetic correlation with other complex traits^35,36^. Heritability was estimated on a liability scale with a sample and population prevalence of 0.5%. Genetic correlations were tested against a comprehensive set of 772 complex traits and diseases. We used within-group corrected *p*-value thresholds to assess the significance of correlation.

## Results and discussion

### Bronchiolitis and asthma

Phenotypes of the 357,869 study individuals, including sex and the mean age during bronchiolitis, are summarized in Supplementary Table S1. We assessed whether severe bronchiolitis requiring hospitalization affects the risk for developing asthma later in life. As shown in Supplementary Table S2, the prevalence of asthma was substantially higher in individuals with bronchiolitis (38,6%) compared to those with no history of LRIs (10,0%). We further detected higher odds of asthma development in individuals with non-RSV bronchiolitis (55,6%) compared to individuals with RSV bronchiolitis (38,6%) (Supplementary Table S2). The findings are consistent with previous reports of asthma after bronchiolitis in adolescence and adulthood^37,38^. Most of the non-RSV bronchiolitis samples comprise cases with unspecified bronchiolitis, and thus may include unrecorded cases of RSV. However, the proportions of RSV and non-RSV are in line with previously observed viral causes of bronchiolitis in Finland in cases under six months, one year, and two years of age (Supplementary table S1)^22,39^. As expected, the median age during the first bronchiolitis was lower in cases with RSV compared to cases with other viruses (Supplementary Table S1). These data indicate that hospitalization due to bronchiolitis during the first two years of life is strongly associated with the risk of asthma during adolescence as well as in later life.

### Genetic loci associated with bronchiolitis in the genome-wide investigation

Genome-wide association (GWA) analysis was performed with 1,465 infants admitted to hospital with bronchiolitis (< 2y) and 356,404 controls without a history of LRIs. The GWAS of non-RSV and RSV bronchiolitis (sub-analysis) were conducted with 484 and 263 cases, respectively, against the same set of controls. The lambda (0.5) values of 1.027, 0.94, and 0.801 in the GWA analyses of bronchiolitis, non-RSV bronchiolitis, and RSV bronchiolitis indicated no genome-wide inflation but suggested that the sub-analysis could be underpowered. The GWAS of bronchiolitis by any causative virus found two associated (*p*<5×10^-8^) loci, located within cadherin related family member 3 (*CDHR3*) and gasdermin B (*GSDMB*) (Figure 1, Table 1).

**Figure 1.**
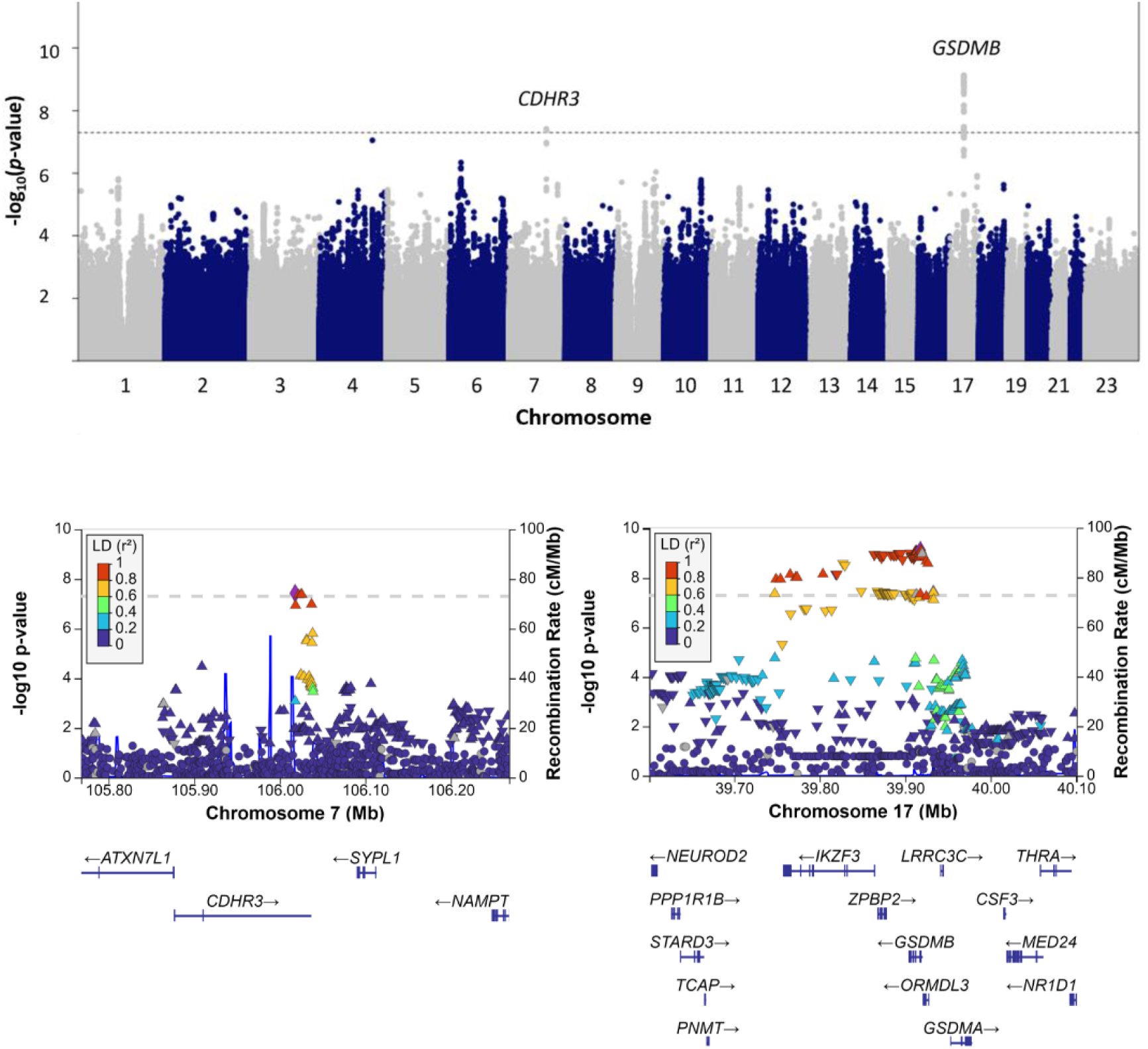
Manhattan plot and regional association plots of the GWAS of bronchiolitis. The y- and x-axes show –log10 *p-*values *vs.* chromosomal positions of the tested variants. The dashed line denotes the threshold of genome-wide significance (*p*<5×10^−8^).

**Table 1.** The loci associated (*p*<5e-8) with bronchiolitis in the GWAS of 1,465 cases and 356,404 controls. The variants with the lowest *p*-values shown.

| Chromosome | Rsid | Position* | Effect allele | Other allele | P-value | Odds ratio | 95% Confidence interval | Frequency | Locus | Most severe annotation |
| --- | --- | --- | --- | --- | --- | --- | --- | --- | --- | --- |
| 7 | rs6967330 | 106018005 | A | G | 3.76e-08 | 1.256 | 1.158-1.363 | 0.268 | CDHR3 | missense variant |
| 17 | rs9303281 | 39917793 | A | G | 7.45e-10 | 1.261 | 1.171-1.358 | 0.448 | GSDMB | noncoding transcript exon variant |

\*Position as base pairs in Grch38 coordinates.

*CDHR3* rs6967330 missense variant with an A to G base change was previously associated with phenotypes including childhood onset asthma^40^, rhinosinusitis^41^, otitis media^42^, and non-RSV bronchiolitis^21^. The transmembrane protein encoded by *CDHR3* is highly expressed in the airway epithelia, and it is a known receptor for rhinovirus C^43^. The rs6967330-A increases the cell surface expression of CDHR3 in the airway epithelium, which is in turn associated with increased replication of rhinovirus-C in the bronchial epithelium^44,45^. In line with the known role of *CDHR3* in rhinovirus pathophysiology, our results found nominal association in the analysis of non-RSV bronchiolitis (*p*=6e-3) but not in the analysis of RSV bronchiolitis (*p*=0.29) (Figure 2A). The effect size was smaller in the RSV compared to overall and non-RSV bronchiolitis, although the differences were not statistically significant, likely due to a small sample size in the sub-analysis (Figure 2A). Taken together, our results suggest that the *CDHR3* variant rs6967330-A increases susceptibility to acute viral bronchiolitis in general, particularly if the causative agent is other than RSV. Studying specific causative viruses of bronchiolitis and their interaction with CDHR3 in molecular biological studies could provide further insight into the pathogenesis of both bronchiolitis and asthma. We suggest that acute viral bronchiolitis by other agents than RSV could act as an environmental trigger towards asthma in genetically susceptible individuals defined by the rs6967330-A genotype.

**Figure 2.**
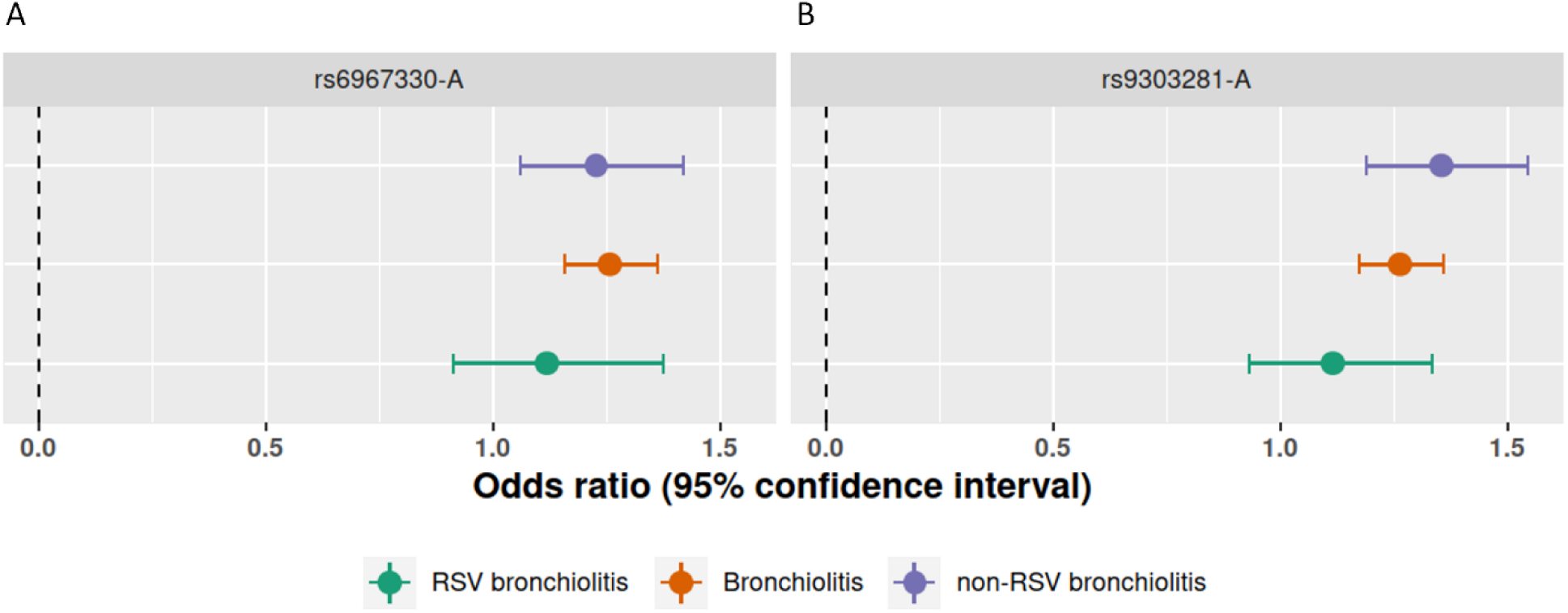
Effect sizes of the associated loci in the GWAS of bronchiolitis and the sub-analysis of RSV and non-RSV bronchiolitis. Effects are shown for the index variant in A) *CDHR3* and B) *GSDMB* locus.

*GSDMB* was previously associated with phenotypes including asthma, childhood-onset asthma, cardiovascular disease, and neutrophil counts in the GWAS Catalog^46^. Variants in the *GSDMB* locus were previously suggested to play a role in rhinovirus but not in RSV wheezing illness in children under three years of age^20^. The current study showed an association with overall bronchiolitis (Table 1, Figure 1), and the sub-analysis found a suggestive association with non-RSV bronchiolitis (rs9303281 *p*=6.2e-6) but no association with RSV bronchiolitis (rs9303281 *p*=0.24). The effect size was largest in the non-RSV bronchiolitis (Figure 2B). Moreover, the non-RSV bronchiolitis effect was significantly larger than the effect in the RSV bronchiolitis GWAS. Thus, we suggest that the *GSDMB* locus mediates the risk of bronchiolitis when the causative virus is not RSV.

### eQTL analysis

There were several hundred associated variants in the *GSDMB* locus in high linkage disequilibrium spanning several genes including *IKZF3*, *ZPBP2*, *ORMDL3*, and *LRRC3C.* Thus, discerning the most likely causal gene is not straightforward. The previous study that found associations with RV wheezing illness at <3y detected that expression levels of *ORMDL3* and of *GSDMB* were increased in rhinovirus-stimulated peripheral blood mononuclear cells (PBMCs), as compared with unstimulated PBMCs^20^. The increase in expression due to exposure to rhinovirus was however not dependent on the genotype. We conducted colocalization analysis to assess if the bronchiolitis-association in the *GSDMB* locus could be mediated by gene expression in other relevant tissues and more specific immune cell types. The analysis suggested that *GSDMB* locus variants were associated with the expression of *GSDMB* and *ORMDL* in cells or tissues including NK cells, B cells, spleen, and CD8 T cells (Supplementary Table S3). Increased gene expression was associated with an increased risk of bronchiolitis in all tested cell types and tissues. Our results suggest that several regulatory variants in the *GSDMB* locus influence *GSDMB* and/or *ORMDL3* expression in distinct immune cell types, and that the changes in gene expression depending on the genotype may mediate susceptibility to severe bronchiolitis.

### Sub-analysis of RSV and non-RSV bronchiolitis

The sub-analysis found suggestively associated (*p*<1e-6) loci with non-RSV and RSV bronchiolitis (Supplementary Table S4). Variant in *SH3D19* showed borderline-significant association with overall bronchiolitis. *SH3D19* encodes SH3 domain containing 19, and the gene was previously associated with traits including height and white blood cell counts in the GWAS Catalog^46^. Interestingly, the gene showed an association with bovine respiratory infections^47^, and suggestive associations with asthma and hospitalization for COVID-19^46^.

### PheWAS

The lead variants of the *CDHR3* and *GSDMB* loci were screened for associations among phenotypes in the FinnGen DF10 (Figure 3, Supplementary Table S5). Both variants showed significant associations with asthma and respiratory system-related phenotypes. Observations with this data further support shared genetic risk factors of asthma and bronchiolitis in the *CDHR3* and *GSDMB* loci.

**Figure 3.**
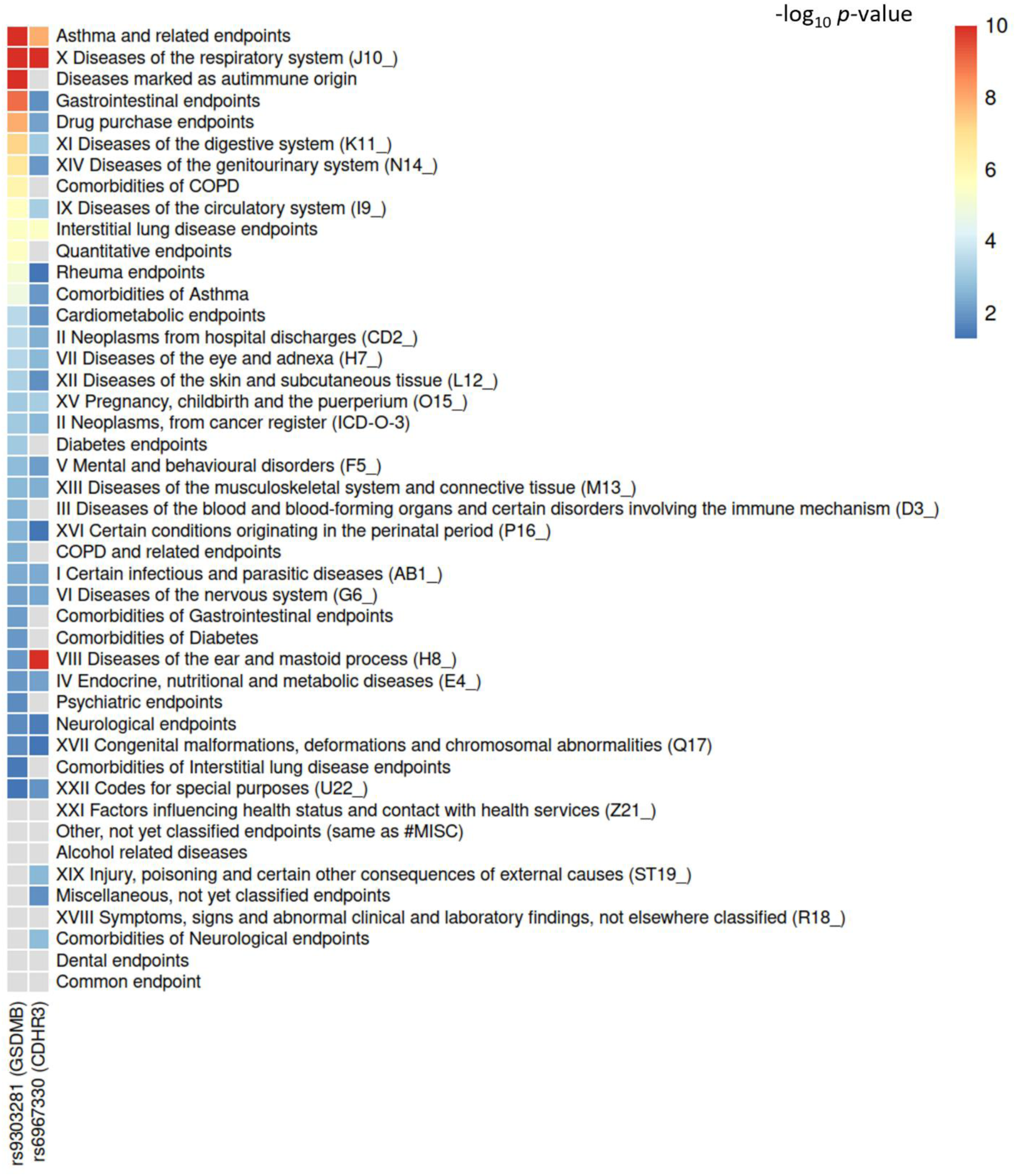
Associations of the bronchiolitis GWAS lead SNPs among the FinnGen DF10 phenotype categories. Association *p*-value is based on the phenotype with the strongest within-category association. The associations are capped into a -log_10_*p*-value of 10.

**Figure 4.**
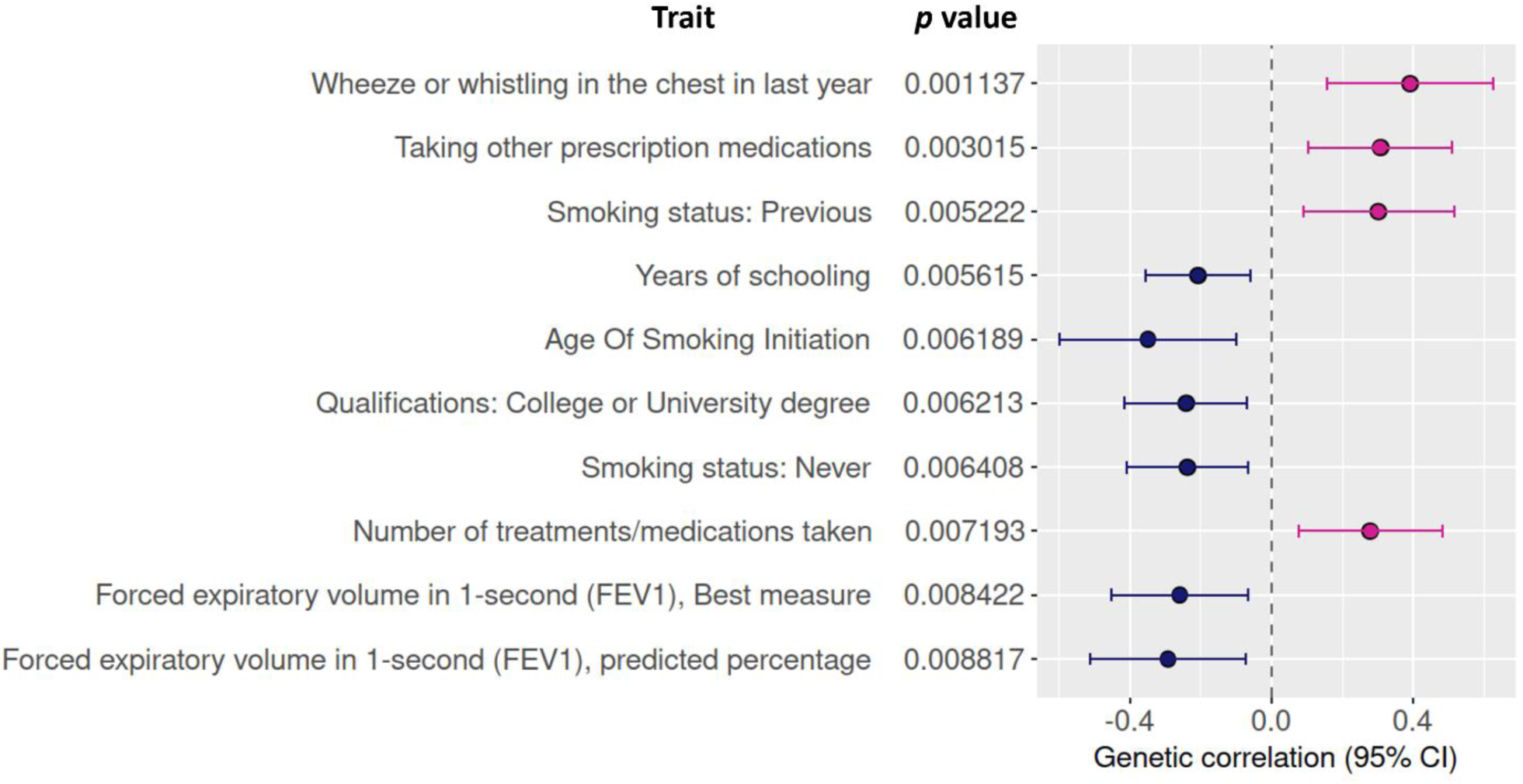
Genetic correlation between bronchiolitis and other complex traits. Genetic correlations were tested in a broad set of 773 complex traits. The figure shows the top 10 correlating traits.

### Genetic correlation and heritability

We used linkage disequilibrium score regression (LDSC) to estimate heritability and shared genetic architecture of bronchiolitis and other complex traits^35^. Wheezing showed a positive correlation with bronchiolitis, and smoking and lung function measures appeared among the top traits with suggestive bronchiolitis-correlations (Figure 3, Supplementary Table S6). Bronchiolitis showed positive correlation with smoking, and negative correlation with never-smoking and spirometric measures of respiratory function. The correlation with the spirometry measures suggests that decreased lung function after bronchiolitis has contributions from genetic factors.

Correlation with smoking could potentially represent an over-the-generation marker of decreased respiratory health mediated by epigenetic changes in the germline due to nicotine consumption^48^. Contrary to conclusions from the two other smoking phenotypes, we detected a negative correlation between bronchiolitis and the low age of smoking initiation. The correlation however presented with large confidence intervals, and the phenotype may not be very informative as it does not specify the duration or magnitude of smoking. Childhood asthma was among the traits that showed genetic correlation with bronchiolitis. The observed genetic correlation highlights the close relationship between bronchiolitis and asthma and shows that this connection has contributions from genetic factors.

The LDSC single nucleotide polymorphism (SNP)-based heritability estimate of bronchiolitis on a total liability scale was 7.6%, with a sample and population prevalences of 0.5%. The estimate suggests that there are further genetic factors of susceptibility to bronchiolitis to be discovered. These could include additional common genetic variants that require a larger number of cases to be detected, and rare genetic variants not captured by the genotyping chips in the current analysis.

In conclusion, *CDHR3* and *GSDMB* loci were associated with overall bronchiolitis in the current GWAS. Supplementary analyses suggested that these loci mediate susceptibility to non-RSV rather than RSV bronchiolitis. We suggest that acute viral bronchiolitis by other causative agents than RSV could trigger asthma in genetically susceptible individuals defined by the *CDHR3* and *GSDMB* genotypes. Based on the importance of non-RSV bronchiolitis in the risk of subsequent asthma, a vaccine protecting against rhinovirus infections in childhood would be of importance. An effective prevention of infant bronchiolitis could potentially have effects on life-long respiratory health by reducing susceptibility to asthma. Moreover, studying specific viral etiologies of bronchiolitis and their interaction with *CDHR3* and *GSDMB* in molecular biological studies could provide further insight into the pathogenesis of both bronchiolitis and asthma.

## Data Availability

GWAS summary data produced in the present study will be available online at the time of the publication.

## Funding

The current study was funded by Stiftelsen Alma och K.A. Snellman Foundation (AP), Sigrid Jusélius Foundation (MH), Foundation for Pediatric Research (MR).

## FinnGen funding statement

The FinnGen project is funded by two grants from Business Finland (HUS 4685/31/2016 and UH 4386/31/2016) and the following industry partners: AbbVie Inc., AstraZeneca UK Ltd, Biogen MA Inc., Bristol Myers Squibb (and Celgene Corporation & Celgene International II Sàrl), Genentech Inc., Merck Sharp & Dohme LCC, Pfizer Inc., GlaxoSmithKline Intellectual Property Development Ltd., Sanofi US Services Inc., Maze Therapeutics Inc., Janssen Biotech Inc, Novartis AG, and Boehringer Ingelheim International GmbH.

## Acknowledgements

We want to acknowledge the participants and investigators of the FinnGen study. CSC-IT Center for Science, Finland, is acknowledged for computational resources. The following biobanks are acknowledged for delivering biobank samples to FinnGen: Auria Biobank (www.auria.fi/biopankki), THL Biobank (www.thl.fi/biobank), Helsinki Biobank (www.helsinginbiopankki.fi), Biobank Borealis of Northern Finland (https://www.ppshp.fi/Tutkimus-ja-opetus/Biopankki/Pages/Biobank-Borealis-briefly-in-English.aspx), Finnish Clinical Biobank Tampere (www.tays.fi/en-US/Research_and_development/Finnish_Clinical_Biobank_Tampere), Biobank of Eastern Finland (www.ita-suomenbiopankki.fi/en), Central Finland Biobank (www.ksshp.fi/fi-FI/Potilaalle/Biopankki), Finnish Red Cross Blood Service Biobank (www.veripalvelu.fi/verenluovutus/biopankkitoiminta), Terveystalo Biobank (www.terveystalo.com/fi/Yritystietoa/Terveystalo-Biopankki/Biopankki/) and Arctic Biobank (https://www.oulu.fi/en/university/faculties-and-units/faculty-medicine/northern-finland-birth-cohorts-and-arctic-biobank). All Finnish Biobanks are members of BBMRI.fi infrastructure (www.bbmri.fi). Finnish Biobank Cooperative -FINBB (https://finbb.fi/) is the coordinator of BBMRI-ERIC operations in Finland. The Finnish biobank data can be accessed through the Fingenious® services (https://site.fingenious.fi/en/) managed by FINBB.

## Abbreviations

GWAS: genome-wide association study
RSV: respiratory syncytial virus
LRI: lower respiratory infection
eQTL: expression quantitative trait locus
PheWAS: phenome-wide association study
LDSC: linkage disequilibrium scire regression

## Notes

### Competing Interest Statement

The authors have declared no competing interest.

### Funding Statement

This study was funded by Stiftelsen Alma och K.A. Snellman Foundation, Sigrid Juselius Foundation, and Foundation for Pediatric Research. FinnGen funding statement: The FinnGen project is funded by two grants from Business Finland (HUS 4685/31/2016 and UH 4386/31/2016) and the following industry partners: AbbVie Inc., AstraZeneca UK Ltd, Biogen MA Inc., Bristol Myers Squibb (and Celgene Corporation & Celgene International II Sarl), Genentech Inc., Merck Sharp & Dohme LCC, Pfizer Inc., GlaxoSmithKline Intellectual Property Development Ltd., Sanofi US Services Inc., Maze Therapeutics Inc., Janssen Biotech Inc, Novartis AG, and Boehringer Ingelheim International GmbH.

### Author Declarations

FinnGen Ethics statement Patients and control subjects in FinnGen provided informed consent for biobank research, based on the Finnish Biobank Act. Alternatively, separate research cohorts, collected prior the Finnish Biobank Act came into effect (in September 2013) and start of FinnGen (August 2017), were collected based on study-specific consents and later transferred to the Finnish biobanks after approval by Fimea (Finnish Medicines Agency), the National Supervisory Authority for Welfare and Health. Recruitment protocols followed the biobank protocols were approved by Fimea. The Coordinating Ethics Committee of the Hospital District of Helsinki and Uusimaa (HUS) statement number for the FinnGen study is Nr HUS/990/2017. The FinnGen study is approved by the Finnish Institute for Health and Welfare (permit numbers: THL/2031/6.02.00/2017, THL/1101/5.05.00/2017, THL/341/6.02.00/2018, THL/2222/6.02.00/2018, THL/283/6.02.00/2019, THL/1721/5.05.00/2019 and THL/1524/5.05.00/2020), Digital and population data service agency (permit numbers: VRK43431/2017-3, VRK/6909/2018-3, VRK/4415/2019-3), the Social Insurance Institution (permit numbers: KELA 58/522/2017, KELA 131/522/2018, KELA 70/522/2019, KELA 98/522/2019, KELA 134/522/2019, KELA 138/522/2019, KELA 2/522/2020, KELA 16/522/2020), Findata permit numbers (THL/2364/14.02/2020, THL/4055/14.06.00/2020, THL/3433/14.06.00/2020, THL/4432/14.06/2020, THL/5189/14.06/2020, THL/5894/14.06.00/2020, THL/6619/14.06.00/2020, THL/209/14.06.00/2021, THL/688/14.06.00/2021, THL/1284/14.06.00/2021, THL/1965/14.06.00/2021, THL/5546/14.02.00/2020, THL/2658/14.06.00/2021, THL/4235/14.06.00/202), Statistics Finland (permit numbers: TK-53-1041-17 and TK/143/07.03.00/2020 (earlier TK-53-90-20) TK/1735/07.03.00/2021, TK/3112/07.03.00/2021) and Finnish Registry for Kidney Diseases permission/extract from the meeting minutes on 4th of July 2019. The Biobank Access Decisions for FinnGen samples and data utilized in FinnGen Data Freeze 9 include: THL Biobank BB2017_55, BB2017_111, BB2018_19, BB_2018_34, BB_2018_67, BB2018_71, BB2019_7, BB2019_8, BB2019_26, BB2020_1, Finnish Red Cross Blood Service Biobank 7.12.2017, Helsinki Biobank HUS/359/2017, HUS/248/2020, Auria Biobank AB17-5154 and amendment #1 (August 17 2020), AB20-5926 and amendment #1 (April 23 2020) and it's modification (Sep 22 2021), Biobank Borealis of Northern Finland_2017_1013, Biobank of Eastern Finland 1186/2018 and amendment 22 /2020, Finnish Clinical Biobank Tampere MH0004 and amendments (21.02.2020 & 06.10.2020), Central Finland Biobank 1-2017, and Terveystalo Biobank STB 2018001 and amendment 25th Aug 2020.

### Summary of Updates

Author affiliations and funding statement were updated.

